# Investigation on the interaction between nifedipine and ritonavir containing antivirus regimens: a physiologically-based pharmacokinetic/pharmacodynamic analysis

**DOI:** 10.1101/2020.05.19.20106658

**Authors:** Wan-jie Niu, Si-ze Li, Sha-sha Jin, Xi-ying Lin, Meng-wan Zhang, Wei-min Cai, Ming-kang Zhong, Xiao-qiang Xiang, Zheng Jiao

## Abstract

**Background and Objective:** Hypertension is a common comorbidity of patients with COVID-19, SARS or HIV infection. Those patients are often treated with commonly used antiviral and antihypertensive agents concomitantly, such as ritonavir-containing regimens and nifedipine. Since ritonavir is a strong inhibitor of CYP3A, when nifedipine is combined with ritonavir-containing antiviral drugs, there is a potential risk of drug-drug interaction. This study aimed to provide guidance on nifedipine treatment during and after co-administration with ritonavir-containing regimens using a physiologically-based pharmacokinetic/pharmacodynamic (PBPK/PD) analysis.

**Methods:** A PBPK/PD model was developed for nifedipine by the software of Simcyp®, and the model was verified using published data. The effects of ritonavir on nifedipine exposures and systolic blood pressure were assessed for instant-release, sustained-release and controlled-release formulations. Moreover, various nifedipine regimens were investigated when co-administrated with and withdrawing ritonavir.

**Results:** PBPK/PD models for three formulations of nifedipine were successfully established. The model predicted pharmacokinetic profiles of nifedipine were comparable to the published data. Ratios of predicted versus observed AU_CDDI_/AUC_Nifedipine_ of nifedipine were within 0.70- to 1.83-fold. Model simulations showed that the inhibitory effect of ritonavir on CYP3A4 increased the C_max_ of nifedipine by 9.82-34.35 times and the AUC_24h_ by 44.94-50.77 times at steady state. Moreover, nifedipine dose reduced to 1/16 of the regular dose during ritonavir co-administration could lead to severe hypotension.

**Conclusions:** Ritonavir had a pronounced influence on the pharmacokinetics and antihypertensive effect of nifedipine. It is not recommended for patients to take nifedipine and ritonavir-containing regimens simultaneously.

## Introduction

The calcium channel blocker (CCB) nifedipine is effective in the treatment of hypertension, angina pectoris and other cardiovascular diseases [1, 2]. European, the United States and Chinese guidelines of hypertension treatment recommend calcium channel blockers (CCB) as the first-line drug therapy [3-5]. The advantages of nifedipine use are rapid onset of action and lack of central nervous system depression. Moreover, comparing to the immediate release (IR) formulation, new once-daily formulations reduce the frequency of nifedipine administration, and thus improve patient compliance. Nifedipine is quickly absorbed after oral administration with peak plasma concentrations occurring in 30 minutes for the IR formulation [6]. It is well absorbed from the gastrointestinal tract, whereas the oral bioavailability of the parent drug is only 45% [7] which suggests that nifedipine undergoes extensive first pass metabolism along the intestine and liver[8, 9]. It is almost completely metabolized by cytochrome P450 (CYP) 3A4 in human body [10]. Therefore, co-administering nifedipine with the strong inhibitors of CYP3A4 may increase its plasma concentrations, leading to the risk of hypotension, hyperglycemia, and conduction disturbances [11].

In recent years, viral infections including Human Immunodeficiency Virus (HIV), Severe Acute Respiratory Syndrome coronavirus (SARS-Cov), Middle East Respiratory Syndrome coronavirus (MERS-Cov), and 2019 novel coronavirus (2019-nCoV) have risen as a global threat to public health. Hypertension, the leading risk factor for mortality worldwide, is a growing problem in viral infections patients [12, 13]. And many virus-infected patients with hypertension are treated with antiviral and antihypertensive agents concomitantly, such as ritonavir (RTV) -containing regimens and nifedipine.

The antiviral drug RTV is a protease inhibitor which can be used as a booster to increase the blood levels of other antiviral medicines including amprenavir, atazanavir, darunavir, fosamprenavir, lopinavir, saquinavir, and tipranavir. Among them, lopinavir/ritonavir (LPV/r) is the most widely used for the treatment of HIV[14] and is regarded as a potential candidate for the treatment of COVID-19 [15-17].

5RTV is a strong time-dependent inhibitor of CYP3A4. Extensive investigations have proved that RTV has significant influence on the pharmacokinetics of CYP3A4 substrates, such as saquinavir[18], quinine[19] and atazanavir[20]. Consequently, coprescription of these drugs leads to the substantial increase of the blood concentration of CYP3A substrates and increase the risk of adverse drug reactions. Regarding the DDI between nifedipine and RTV, the U.S. Food and Drug Administration (FDA)-approved label states that caution is warranted, clinical monitoring of patients is recommended and a dose decrease may be needed for nifedipine when co-administered with RTV[21]. But the detailed guidance is not provided. Therefore, there is an urgent need to address how to adjust the dose of nifedipine when co-administered with RTV-containing regimens especially during the COVID-19 pandemic.

To our knowledge, there have been no reports on DDI between nifedipine and RTV except for a case report[22] which showed that co-administration of nifedipine with RTV may significantly increase nifedipine exposure, leading to severe hypotension and renal failure in a patient with HIV. Therefore, it is risky and costly to investigate the DDI between nifedipine and RTV through traditional clinical trials. While physiologically-based pharmacokinetic (PBPK) models can be of great value for the assessment of the various dose regimens and analysis of the dynamic change in plasma concentrations over time for the victim and inhibitor drugs, and the exploration of the magnitude of DDI[16]. Moreover, PBPK models could link with pharmacodynamic (PD) models to predict changes in drug effect due to extrinsic or intrinsic factors that affect the drug PK[23].

The purpose of this study is to investigate the effects of RTV on nifedipine exposure and systolic blood pressure (SBP) via a PBPK/PD modeling approach. And to apply the model to assess various nifedipine dose regimens in the presence or absence of RTV in order to design the regimen for nifedipine during and after co-administration with RTV-containing therapies.

## Methods

### Nifedipine PBPK model development

The PBPK model for nifedipine was built in Simcyp^®^ simulator (version 16, Certara Inc., Princeton, New Jersey, USA). Nifedipine is a Biopharmaceutics Classification System Class II drug, with low solubility and high intestinal permeability [24]. It is predominantly eliminated through CYP3A4 metabolism[24, 25]. Nifedipine physicochemical properties (LogP and pKa), absorption, distribution, metabolism and elimination (ADME) parameters are summarized in **Table 1**.

**Table 1.** The parameters included in the PBPK model for nifedipine.

| Parameter | Units | Values |
| --- | --- | --- |
| molecular weight | g/mol | 346.3* |
| LogP | - | 2.69* |
| pKa | - | 2.82* (monoprotic base) |
| $f_u$ | - | 0.039* |
| B/P ratio | - | 0.685* |
| **Absorption model** |  | ADAM (SR, CR)<br>1st order(IR) |
| MDCK II permeability | cm/s | $61 \times 10^{-6}$ (IR,SR)[26] |
| $P_{\text{eff,man}}$ in colon | cm/s | $0.17 \times 10^{-4}$ (CR)[27] |
| $f_a$ | - | 1* (IR) |
| $k_a$ | 1/h | 3.67* (IR) |
| **Distribution model** |  | Minimal PBPK |
| $V_{ss}$ | L/kg | 0.57* |
| **Elimination** |  |  |
| rCYP3A4 |  |  |
| $K_m$ | $\mu\text{M}$ | 10.95* |
| $V_{\text{max}}$ | pmol/min/pmol CYP | 22* |
| rCYP3A5 |  |  |
| $K_m$ | $\mu\text{M}$ | 31.9* |
| $V_{\text{max}}$ | pmol/min/pmol CYP | 3.5* |

\*: obtained by SimCYP® simulator (version 16)
ADAM, the Advanced Dissolution, Absorption and Metabolism model; *B/P ratio*, blood to plasma concentration ratio; *CR*, controlled release; ; $f_a$ , fraction available from dosage form; $f_u$ , the fraction unbound in plasma; *IR*, immediate release; $k_a$ , absorption rate constant; $K_m$ , the Michaelis-Menten constant; *LogP*, octanol–water partition; *Minimal PBPK*, Minimal Physiologically-based Pharmacokinetic model; $P_{\text{eff,man}}$ , the effective permeability in humans; *SR*, sustained release; $V_{\text{max}}$ , the maximal enzyme velocity; $V_{ss}$ , volume of distribution at steady-state.

The PK profiles for nifedipine were predicted using the Simcyp^®^ nifedipine compound file, with a minimal PBPK distribution model and elimination pathway characterized by enzyme kinetics. There are three available nifedipine formulations on the market, namely IR, sustained-release (SR) and controlled-release (CR) formulation. This study investigated all three formulations. The first order model was used to describe the absorption process of nifedipine IR. For the SR and CR nifedipine, the oral absorption was described by the Advanced Dissolution, Absorption and Metabolism (ADAM) model within Simcyp^®^ using *in vitro* dissolution data. Dissolution data of the CR and SR formulation were obtained from literature [27] and package insert of Adalat^®^[28], respectively.

### Nifedipine PBPK model verification

**IR formulation** PBPK model of nifedipine IR was verified using clinical DDI data with CYP3A inhibitors/inducers. The predictive performance of DDI for nifedipine PBPK model was investigated by using the perpetrators of diltiazem and rifampicin, which all have the built in compound files in Simcyp^®^. Observed data of nifedipine PK from six published DDI studies [1, 2, 29-32] was captured using GetData Graph Digitizer (version 2.22, www.getdata-graph-digitizer.com). Details of these clinical DDI studies were summarized in **Table 2**.

**Table 2.** Summary of published clinical DDI studies used for verifying the nifedipine PBPK model.

| Study | Route of administration | Dose | Information | References |
| --- | --- | --- | --- | --- |
| **CYP3A inhibitors** |  |  |  |  |
| diltiazem | oral | nifedipine 20 mg + diltiazem 60 mg | Effect of duration of diltiazem pretreatment on nifedipine kinetics | Ohashi et al. [29] |
|  | oral | I. nifedipine 20 mg + diltiazem 30 mg<br>II. nifedipine 20 mg + diltiazem 90 mg | Dose dependent effect of diltiazem on the PK of nifedipine | Tateishi et al.[30] |
|  | oral | diltiazem 60 mg TID ×3 days + nifedipine 20 mg | Effects of diltiazem on the PK of nifedipine | Ohashi et al. [31] |
| **CYP3A inducer** |  |  |  |  |
| rifampicin | oral<br>IV | rifampicin 600 mg QD 7 days + nifedipine 20 mg<br>rifampicin 600 mg QD 7 days + nifedipine 20 µg/kg | Nifedipine-rifampin interaction | Holtbecker et al.[32] |
|  | oral | rifampicin 1200 mg+ nifedipine 10 mg (administered 8 h after treatment of rifampicin) | Effect of single dose of rifampicin on the PK of nifedipine | Ndanusa et al. [1] |

*TID*, three times a day; *Q8H*, every 8 hours; *QD*, once daily; *IV*, intravenous; *PK*, pharmacokinetic; *PD*, pharmacodynamics

**SR and CR formulations** The PBPK models for SR and CR formulations were also verified with PK data from a single-dose administration in healthy volunteers [33, 34]. The predicted area under the plasma concentration-time curve (AUC) ratios of nifedipine were compared with the respective observations.

The accuracy of prediction was measured by calculating the fold error between predicted and observed, described as (Eq.1). C_max_ and AUC ratios were estimated for all three formulations. Evaluation criteria is the ratio of predicted AUC and C_max_ values are within 2-fold namely, 0.5≤ratio≤2.0 of the observed values.

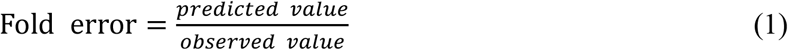

### Nifedipine-RTV DDI prediction

First, the changes of CYP3A4 abundance in liver and intestine with or without RTV were predicted in order to explore the inhibition of RTV on CYP3A4.

Then, the established nifedipine PBPK model was used to predict multiple dosing of nifedipine when co-administered with RTV-containing regimens. Since RTV is the only clinical inhibitor and inducer of CYP3A4 within the regimen, only RTV was simulated as a surrogate for the RTV-containing regimens [35]. In addition, the PBPK model of RTV has been already verified by simulating its inhibition effects on the PK profiles of CYP3A4 substrates [36]. Thus, the RTV model was not herein verified.

IR nifedipine at 10 mg with repeated dose administration every 12 hours (Q12H)alone was simulated to reach the steady state, followed by the combination of 100 mg Q12H RTV and 10 mg Q12H nifedipine for 14 days. The commonly used regimen for SR and CR formulations 30 mg Q12H and 60 mg every 24 hours (Q24H), respectively were also simulated. Considering no consensus for the use of nifedipine in patients receiving RTV, PK profiles at different dose levels of nifedipine was simulated to investigate the optimal dose during the co-administration with RTV.

After RTV was discontinued, 2 different dose regimens were investigated. One is to use reduced-dose nifedipine, and the other is to restore to the original dose of nifedipine. Taking IR tablets as an example, in the first regimen, nifedipine at a lower dose (5 mg Q12H) or an extended dosing interval (10 mg Q24H) was taken for 5 more days after the last dose of RTV, followed by a return to an original regimen (10 mg Q12H). Moreover, lower dose of nifedipine (1.25 mg, 1/4 of the minimum specification of IR nifedipine tablets) was investigated. Since chewing or crushing before swallowing SR or CR tablets is not allowed, dose less than 30 mg or 60 mg was not assessed. In the second regimen, the regular dose of nifedipine (10 mg Q12H) was taken immediately after the last dose of RTV. The detailed dose adjustment scenarios are shown in **Table 3**. Due to the limitation of software, the dose amount of SR and CR tablets cannot be adjusted directly and only the adjustment of the dosing interval was assessed.

**Table 3.** Clinical scenarios used in the nifedipine PBPK model simulation.

| Nifedipine formulation | Combined with RTV | After withdrawal of RTV |  |
| --- | --- | --- | --- |
|  |  | Nifedipine regimen 1 | Nifedipine regimen 2 |
| Immediate Release (IR) | I. 10 mg Q12H × 3 days + (5 mg Q12H + RTV 100 mg Q12H) × 14 days | 5 mg Q12H × 5 days + 10 mg Q12H | 10 mg Q12H |
| | II. 10 mg Q12H × 3 days + (10 mg Q24H + RTV 100 mg Q12H) × 14 days | 10 mg Q24H × 5 days + 10 mg Q12H | 10 mg Q12H |
| | III. 10 mg Q12H × 3 days + (1.25 mg Q12H + RTV 100 mg Q12H) × 14 days | 1.25 mg Q12H × 5 days + 10 mg Q12H | 10 mg Q12H |
| | IV. 10 mg Q12H × 3 days + (1.25 mg Q24H + RTV 100 mg Q12H) × 14 days | 1.25 mg Q24H × 5 days + 10 mg Q12H | 10 mg Q12H |
| Sustained Release (SR) | 30 mg Q12H × 3 days + (30 mg Q24H + RTV 100 mg Q12H) × 14 days | 30 mg Q24H × 5 days + 30 mg Q12H | 20 mg Q12H |
| Controlled Release (CR) | 60 mg Q24H × 3 days + (60 mg Q48H + RTV 100 mg Q12H) × 14 days | 60 mg Q48H × 5 days + 60 mg Q24H | 60 mg Q24H |

*Q12H*, every 12 hours; *Q24H*, every 24 hours; *Q48H*, every 48 hours; *RTV*, ritonavir.

### Nifedipine PD model development and verification

Studies have demonstrated that it is more important to control systolic blood pressure (SBP) than diastolic blood pressure (DBP)[37], and SBP is a better predictor of cardiovascular risk than DBP in most of patients treated with antihypertensive agents[38]. Therefore, only SBP was employed in the PBPK/PD modeling. A E_max_ model developed by Shimada *et al*. [39] was linked to the PBPK to investigate the effect of DDI on the SBP. The relationship between nifedipine concentration and the reduction in SBP was expressed by ordinary E_max_ model (Eq. 2).

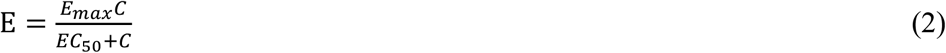

where E, *E*_max_*, EC*_50_ *and C* represent the reduction in SBP, the maximum reduction in SBP, nifedipine concentration at 50% maximum effect and nifedipine concentration, respectively.

The E_max_ and the EC_50_ were -35 mmHg and 35 nM for therapeutic dose in the previous report, respectively[39]. However, in reported cases of nifedipine overdose, the SBP of the patients would decrease about 50 mmHg [6, 40, 41]. Therefore, in this study, the E_max_ value was increased to -50 mmHg to fit the maximal SBP decrease based on the previous case reports of nifedipine overdose. In addition, based on the range of EC_50_ values reported in the literature [39, 42], PD model under different EC_50_ values was examined to fit the observed SBP change caused by nifedipine at therapeutics doses [43, 44]. Details of these clinical PD studies were summarized in **Table 4**. The accuracy of prediction was measured by comparing the maximum reduction in SBP (R_max_) and the area under the effect-time curve (AUE) between prediction and observation. The acceptable criteria were within 2-fold error.

**Table 4.**
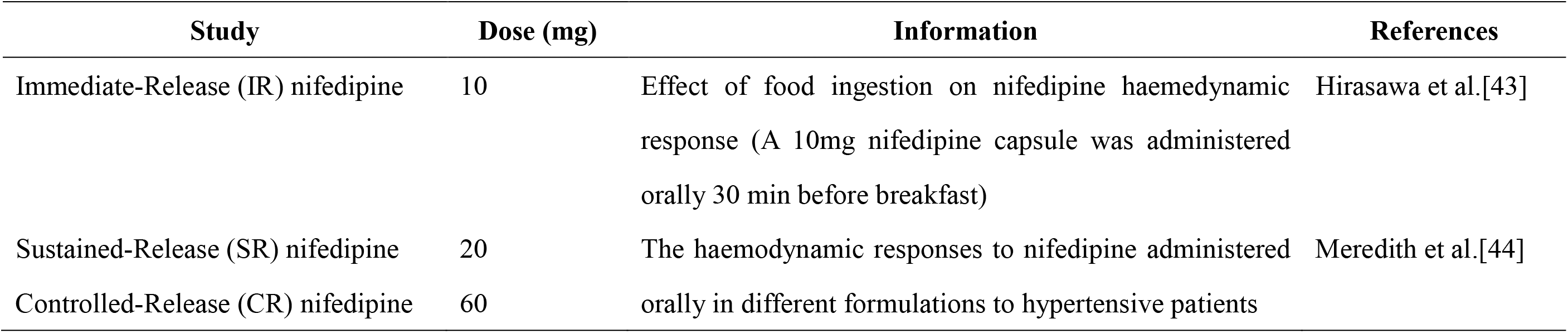
Summary of published clinical PD studies used for verifying the nifedipine PD model.

| Study | Dose (mg) | Information | References |
| --- | --- | --- | --- |
| Immediate-Release (IR) nifedipine | 10 | Effect of food ingestion on nifedipine haemodynamic response (A 10mg nifedipine capsule was administered orally 30 min before breakfast) | Hirasawa et al.[43] |
| Sustained-Release (SR) nifedipine | 20 | The haemodynamic responses to nifedipine administered orally in different formulations to hypertensive patients | Meredith et al.[44] |
| Controlled-Release (CR) nifedipine | 60 |  |  |

### Nifedipine PBPK/PD model application

The developed PBPK/PD model was used to predict the changes in SBP caused by the dynamic changes in nifedipine exposures with and without RTV. Two clinical scenarios were simulated and all the model simulations were carried out using a virtual population representative within Simcyp^®^.

The first scenario was designed to answer whether dose adjustment of nifedipine can maintain SBP at normal range. Two dosing regimens were investigated after patients taking combined nifedipine with RTV at steady state, (1) the nifedipine dose was continued at regular regimen during co-administration with RTV; (2) the nifedipine regimen was changed to extended dosing interval during the co-administration with RTV.

The second scenario was designed to investigate how to adjust the dose regimen of nifedipine after the withdrawal of RTV. For this scenario, two dosing regimens were examined, (1) adjusted dosage regimen was continued for 5 more days after the last dose of RTV; (2) the regular regimen was resumed immediately after RTV was stopped.

## Results

### Development of the nifedipine PBPK model

The model of IR nifedipine was developed first using the Simcyp^®^ compound file. Next was development of the SR and CR nifedipine model which used the same input data as the IR model except for the absorption model and dissolution data. The IR nifedipine used the first-order absorption model, while the SR and CR formulations used the ADAM model.

### Verification of the nifedipine PBPK model

Concentration–time plasma profiles from DDI studies were used to verify the PK model of nifedipine. The PBPK model predictions of nifedipine plasma concentrations profiles in three formulations (IR/SR/CR) were consistent with the clinically observed data and met the model acceptance criteria. The detailed results are as follows:

**IR formulation** Using 6 published DDI studies[1, 2, 29-32], the predicted AUC and C_max_ ratios of nifedipine after single or multiple administrations in the presence and absence of concomitant drugs were compared with the respective observations (**Table 5; Figure 1-2**). The results show that the predicted nifedipine AUC values in the absence of concomitant drugs were consistent with observed data, and median fold-error was 0.66 (range: 0.29-1.84). In the presence of concomitant drugs, the predicted AUC_DDI_/AUC_Nifedipine_ value was within 0.70-1.83-fold of the observed AUC_DDI_/AUC_Nifedipine_ value. Relatively good predictability of the C_max_ ratios within 0.52-fold was also confirmed, in comparison with the respective clinical observations. Thus, the current nifedipine model demonstrated good performance for the purpose of DDI investigation.

**Table 5.**
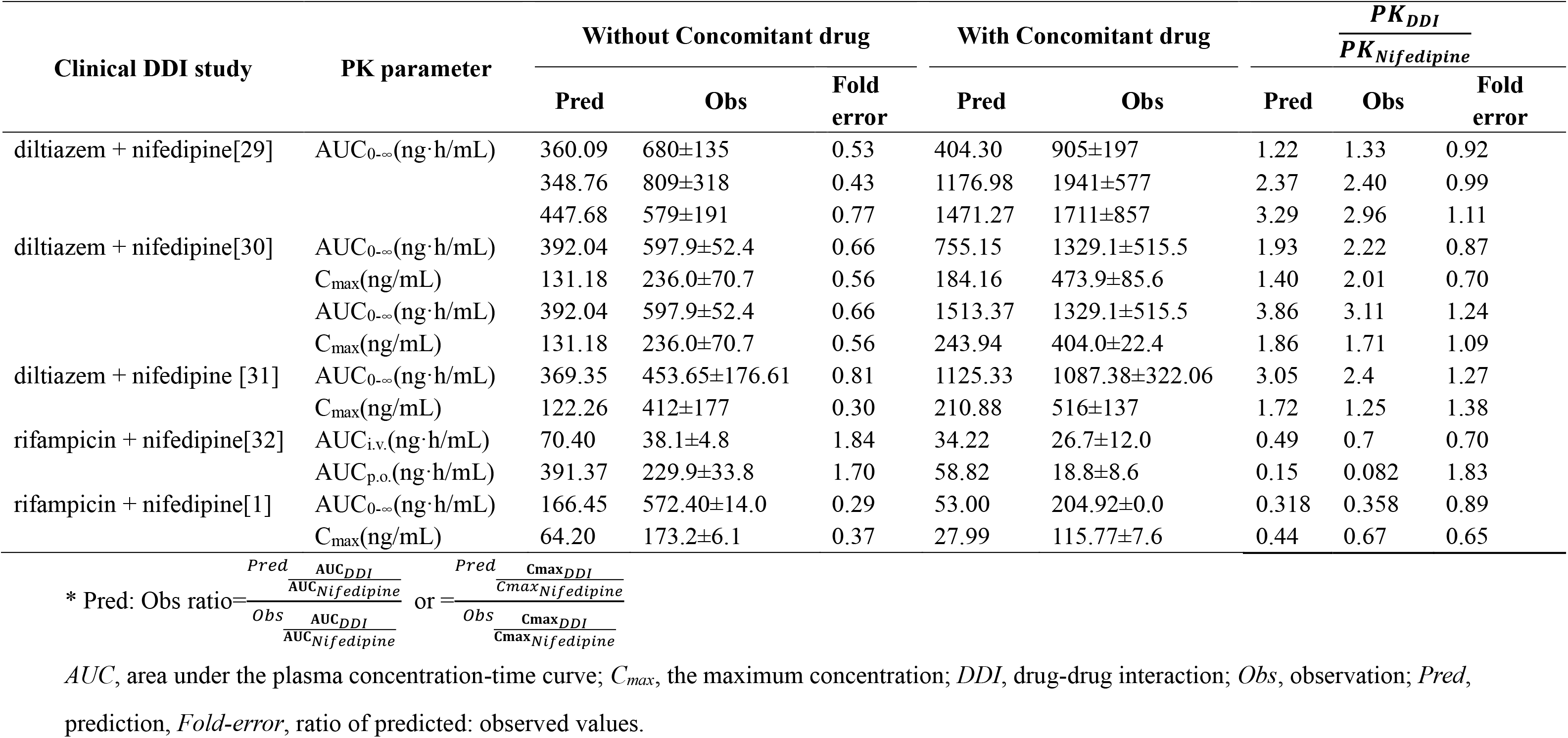
Comparison of IR formulation PBPK model predicted and clinically observed pharmacokinetic parameter.

**Figure 1.**
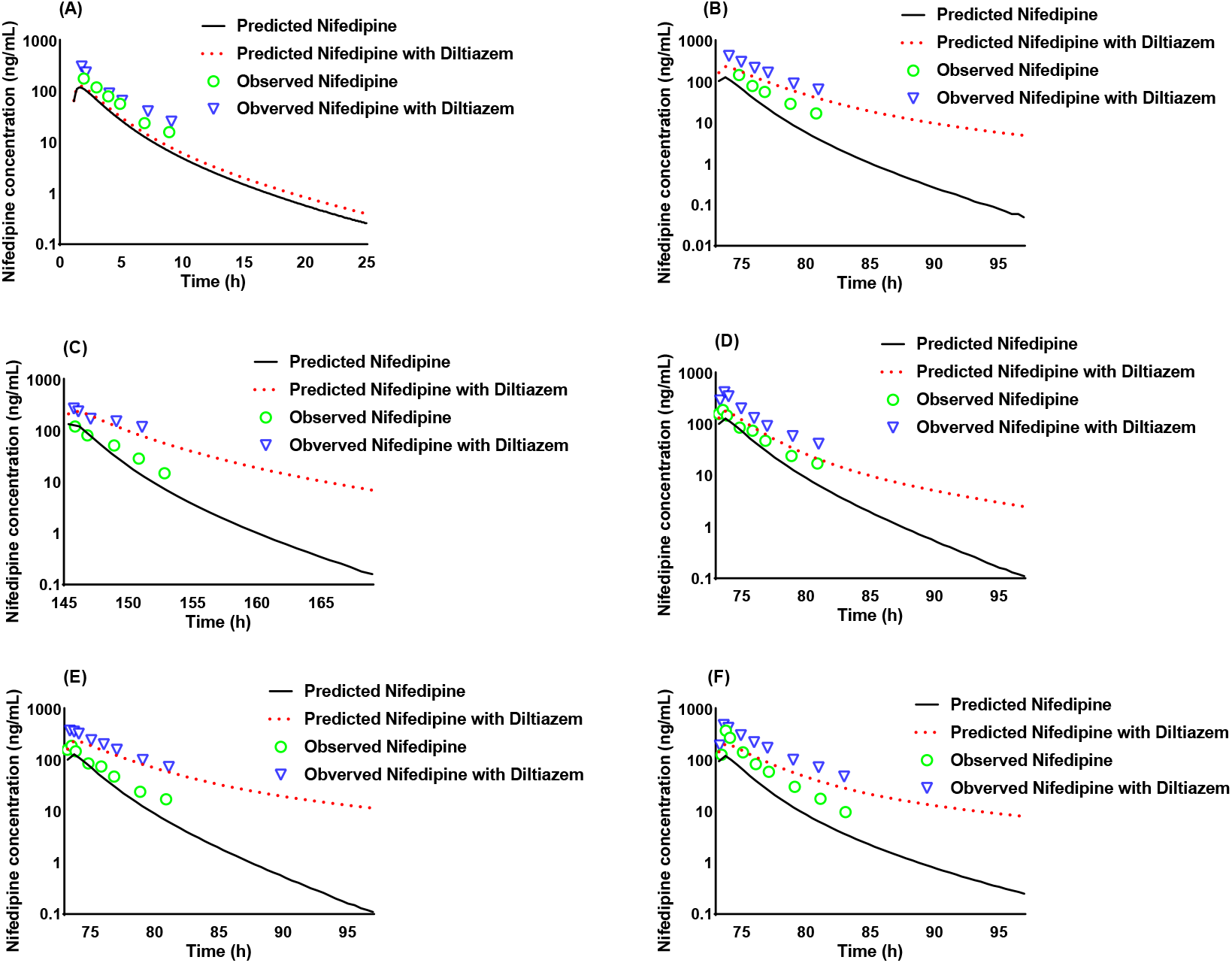
Comparison of PBPK model predictions of plasma concentrations of IR formulation nifedipine. Panel A-F give predicted PK profiles of nifedipine in the presence (red dotted lines) and absence (balck lines) of diltiazem. The green circles and blue triangles represented the observed nifedipine concentration in the presence and absence of concomitant drugs, respectively. (A) nifedipine 20 mg + diltiazem 60 mg; (B) nifedipine 20 mg + diltiazem 60 mg Q8H×10 doses; (C) nifedipine 20 mg + diltiazem 60 mg Q8H×19 doses; (D) nifedipine 20 mg + diltiazem 30 mg; (E) nifedipine 20 mg + diltiazem 90 mg; (F) nifedipine 20 mg + diltiazem 60 mg TID ×3 days.

**Figure 2.**
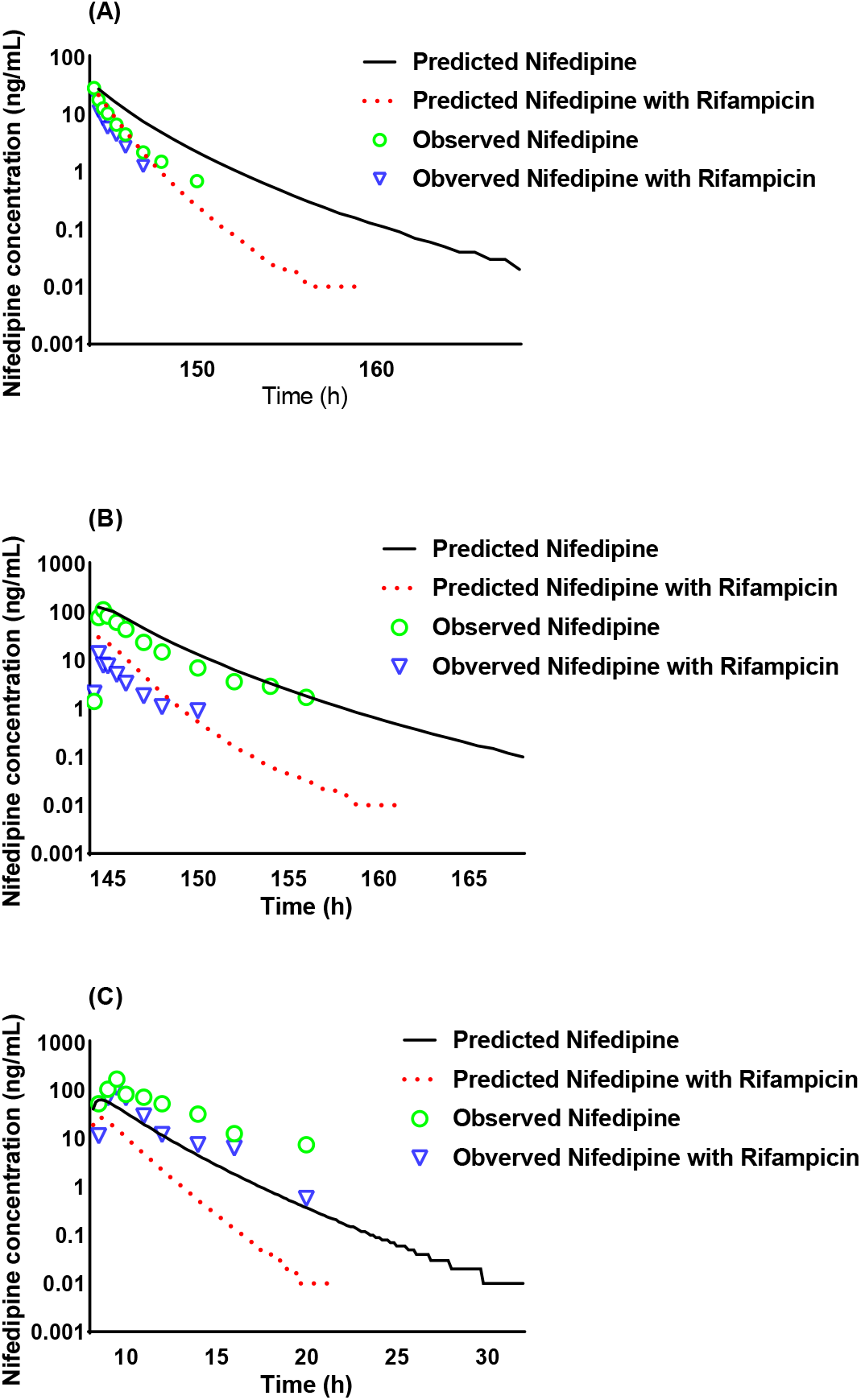
Comparison of PBPK model predictions of plasma concentrations of IR formulation nifedipine. Panel A-C give predicted PK profiles of nifedipine in the presence (red dotted lines) and absence (black lines) of rifampicin. The green circles and blue triangles represented the observed nifedipine concentration in the presence and absence of concomitant drugs, respectively. (A) rifampicin 600 mg QD 7 days + nifedipine 20 μg/kg, infusion; (B) rifampicin 600 mg QD 7 days + nifedipine 20 mg, oral; (C rifampicin 1200 mg, nifedipine 10 mg (administered 8 h after pre-treatment of rifampicin.).

**SR formulation** As shown in **Figure 3**, the PBPK model for SR formulation resulted in a good agreement between observed and predicted values for nifedipine PK profiles after single oral dose administration in the healthy volunteers. And thepredicted AUC ratio was 1.45-fold of the observed AUC which indicated good predictive performance.

**Figure 3.**
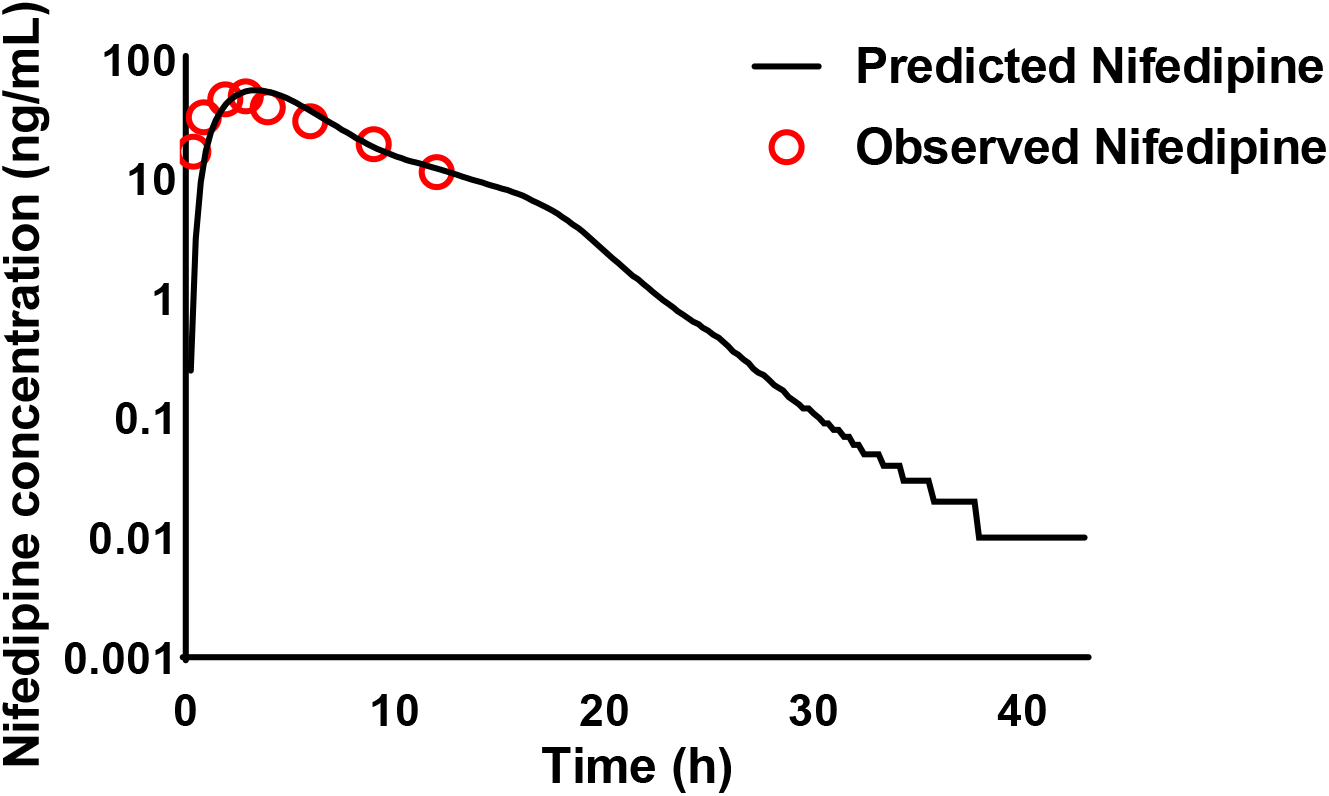
Comparison of physiologically-based pharmacokinetic (PBPK) model predictions (black lines) of plasma concentrations of SR formulation nifedipine in healthy volunteers with 20 mg single dose. Clinical data is represented as red circles.

**CR formulation Figure 4** shows the observed and predicted values for CR nifedipine PK profiles after a single oral dose administration in the healthy volunteers. The predicted C_max_ and AUC ratios were within 1.46-fold of the observed data. Therefore, the PBPK model for CR formation showed a good descriptive and predictive performance.

**Figure 4.**
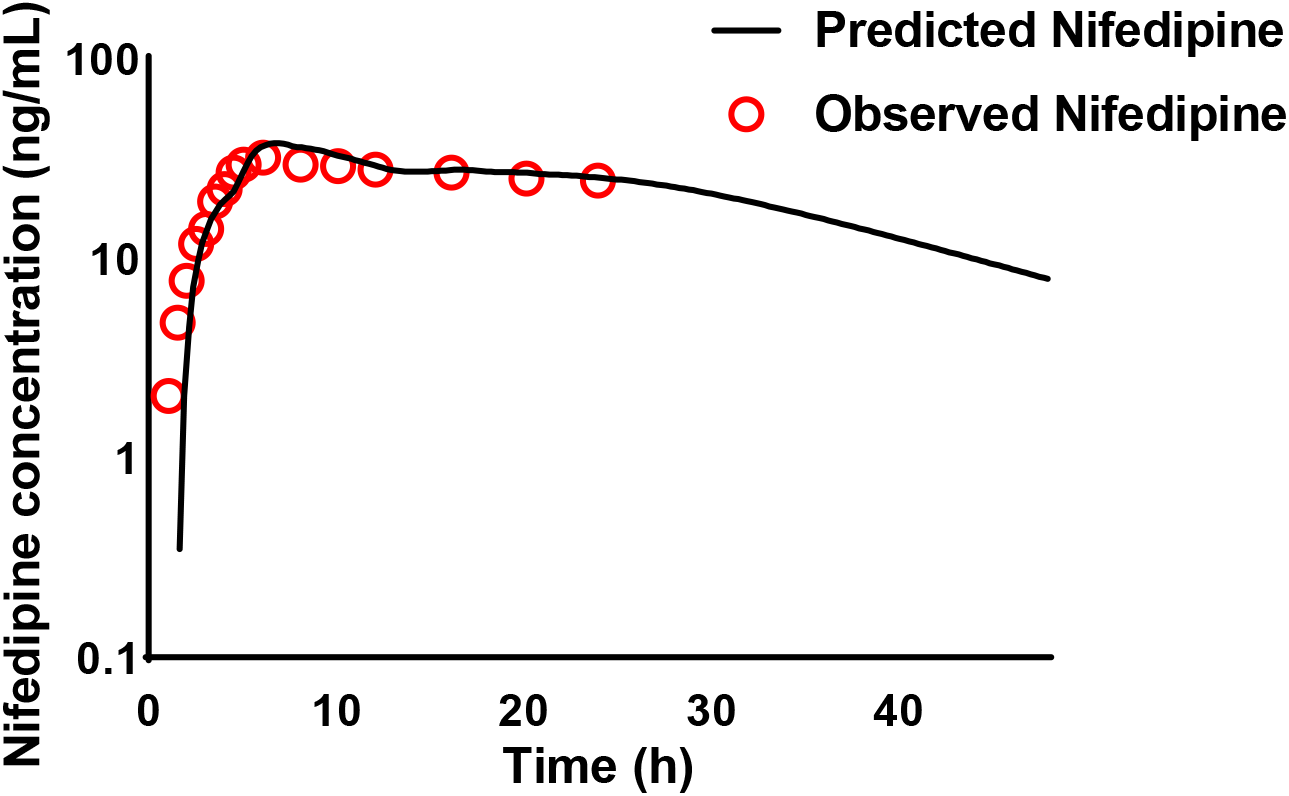
Comparison of physiologically-based pharmacokinetic (PBPK) model predictions (black lines) of plasma concentrations of CR formulation nifedipine in healthy volunteers after a 60 mg oral dose. Clinical data is represented as red circles.

### Nifedipine-RTV DDI

The final PBPK model was used to predict changes in CYP3A4 enzyme abundance and nifedipine plasma PK profiles after the co-administration with RTV over time. As shown in **Figure 5**, the CYP3A4 in the liver and intestinal were maximally deactivated within three days after the co-administration of RTV (100 mg Q12H), which suggested there was a strong DDI between nifedipine and RTV **Figure 6** shows the predicted PK profiles of three formulations nifedipine over time following the dosing schedules listed in **Table 3**. Both the C_max_ and AUC increased significantly due to CYP3A4 inhibition by RTV and reached steady state on approximately Day 10. The inhibitory effect of RTV on CYP3A4 increased the C_max_ of nifedipine by 9.82-34.35 times and the AUC24h by 44.94-50.77 times (**Table 6**), which showed that the combination of RTV have a significant impact on the exposure of nifedipine.

**Table 6.** A summary of PK parameters of nifedipine after co-administration with RTV using the final PBPK model.

| Parameter | IR formulation |  | SR formulation |  | CR formulation |  |
| --- | --- | --- | --- | --- | --- | --- |
|  | nifedipine | nifedipine + RTV | nifedipine | nifedipine + RTV | nifedipine | nifedipine + RTV |
| $T_{\max}$ (h) | 4.14 | 8.15 | 8.57 | 12.23 | 8.34 | 9.68 |
| $C_{\max}$ (ng/mL) | 65.79 | 745.86 | 42.11 | 1241.12 | 41.48 | 1683.39 |
| $AUC_{24h}$ (ng/mL·h) | 275.80 | 16053.90 | 508.03 | 28109.70 | 694.79 | 38534.48 |
| $C_{\max}$ Ratio ($\frac{nifedipine + RTV}{nifedipine}$) | | 9.82 | | 28.71 | | 34.35 |
| $AUC$ Ratio ($\frac{nifedipine + RTV}{nifedipine}$) | | 50.77 | | 49.58 | | 44.94 |

$AUC$ , area under the plasma concentration-time curve; $C_{\max}$ , the maximum concentration; $CR$ , controlled release; $IR$ , immediate release; $RTV$ , ritonavir; $SR$ , sustained release; $T_{\max}$ , time to reach maximum plasma concentration.

**Figure 5.**
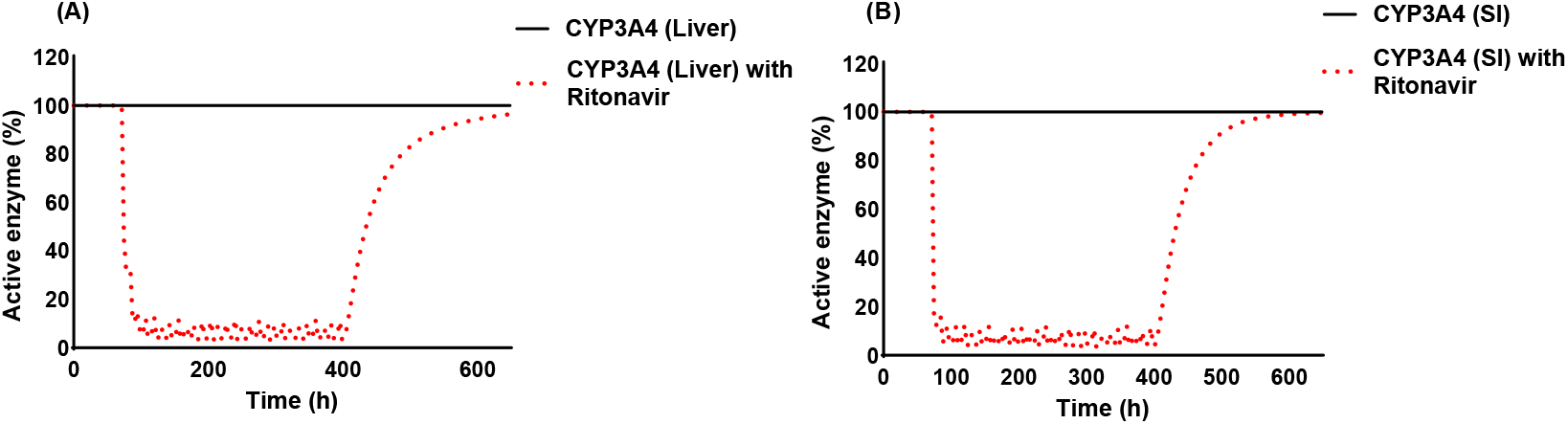
Predictions of active CYP3A4 abundance profiles in (A) liver and (B) intestinal over time after multiple oral administration of ritonavir. The red and black solid lines represent the concentration curves of combined nifedipine with RTV and nifedipine alone, respectively.

**Figure 6.**
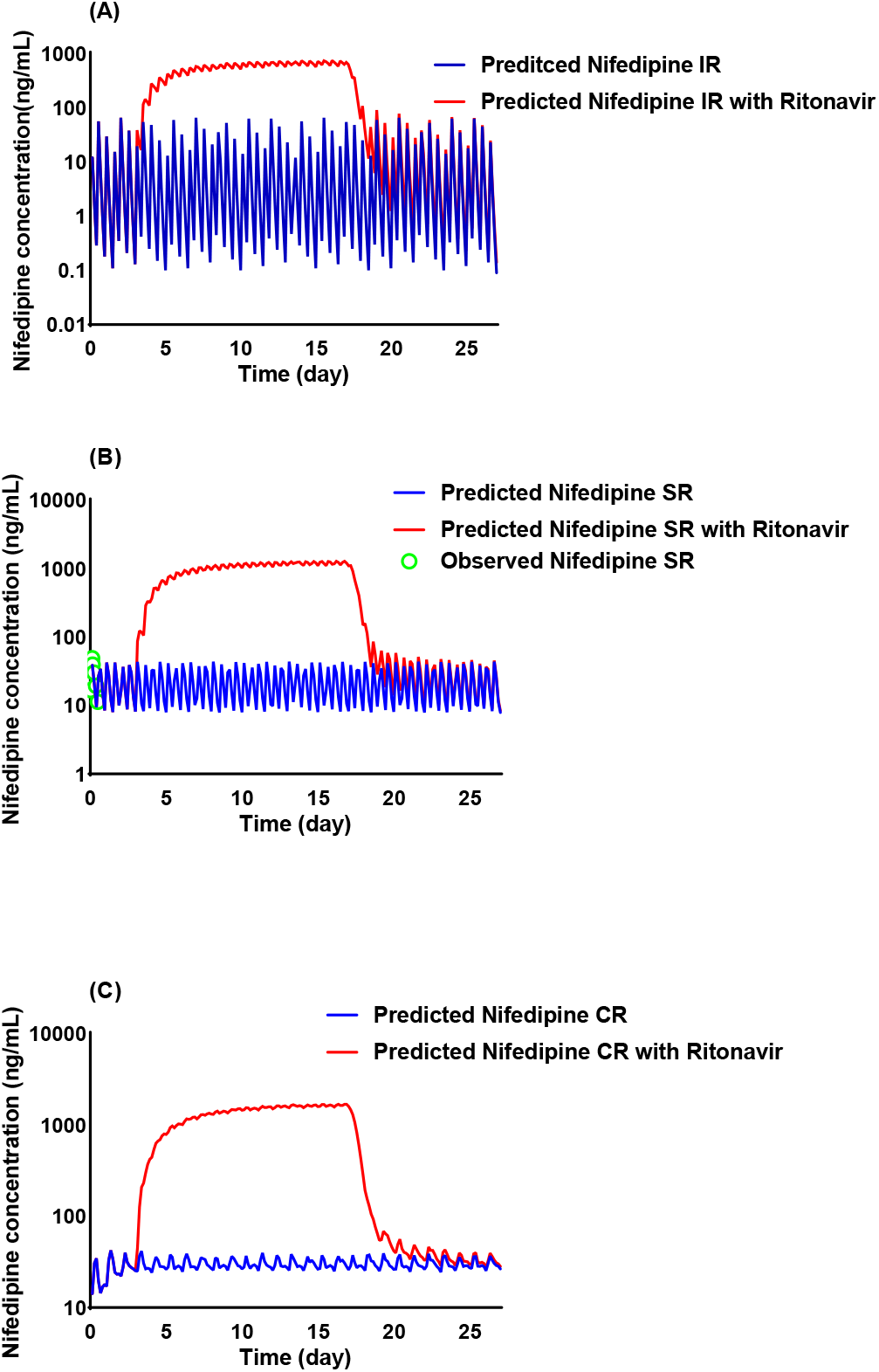
Model prediction of time-based changes in the drug–drug interaction (DDI) magnitude of nifedipine and ritonavir (RTV) over multiple days. (A) IR nifedipine 10 mg every 12 hours (Q12H) × 3 days + (nifedipine 10 mg Q12H + RTV 100 mg Q12H) × 14days+ nifedipine 10 mg Q12H × 10 days. (B) SR nifedipine 30 mg Q12H × 3 days + (30 mg Q12H +RTV 100 mg Q12H) × 14 days+ nifedipine 30 mg Q12H × 10 days. (C) CR nifedipine 60 mg Q24H × 3 days+ (60 mg Q24H + RTV 100 mg Q12H) × 14 days+ nifedipine 60 mg Q24H × 10 days. The red and blue solid lines represent the concentration curves of combined nifedipine with RTV and nifedipine alone, respectively.

Moreover, the plasma concentration of three formulations of nifedipine decreased to the baseline (without RTV) on the 4-5^th^ day after the last dose of RTV

### Verification of the nifedipine PD model

When the EC_50_ was set to 98 nM, the PD model fitted best. The model-predicted SBP compared with the observations for nifedipine at regular dose were presented in **Figure 7**. And the predicted PD profiles for three formulations nifedipine at regular doses suggested that the model was successful in predicting the clinical data. The ratios of the predicted and observed values of R_max_ and AUE for IR, SR and CR nifedipine were within 0.87-1.14 (**Table 7**). Thus, the current nifedipine PD model showed good predictive performance.

**Figure 7.**
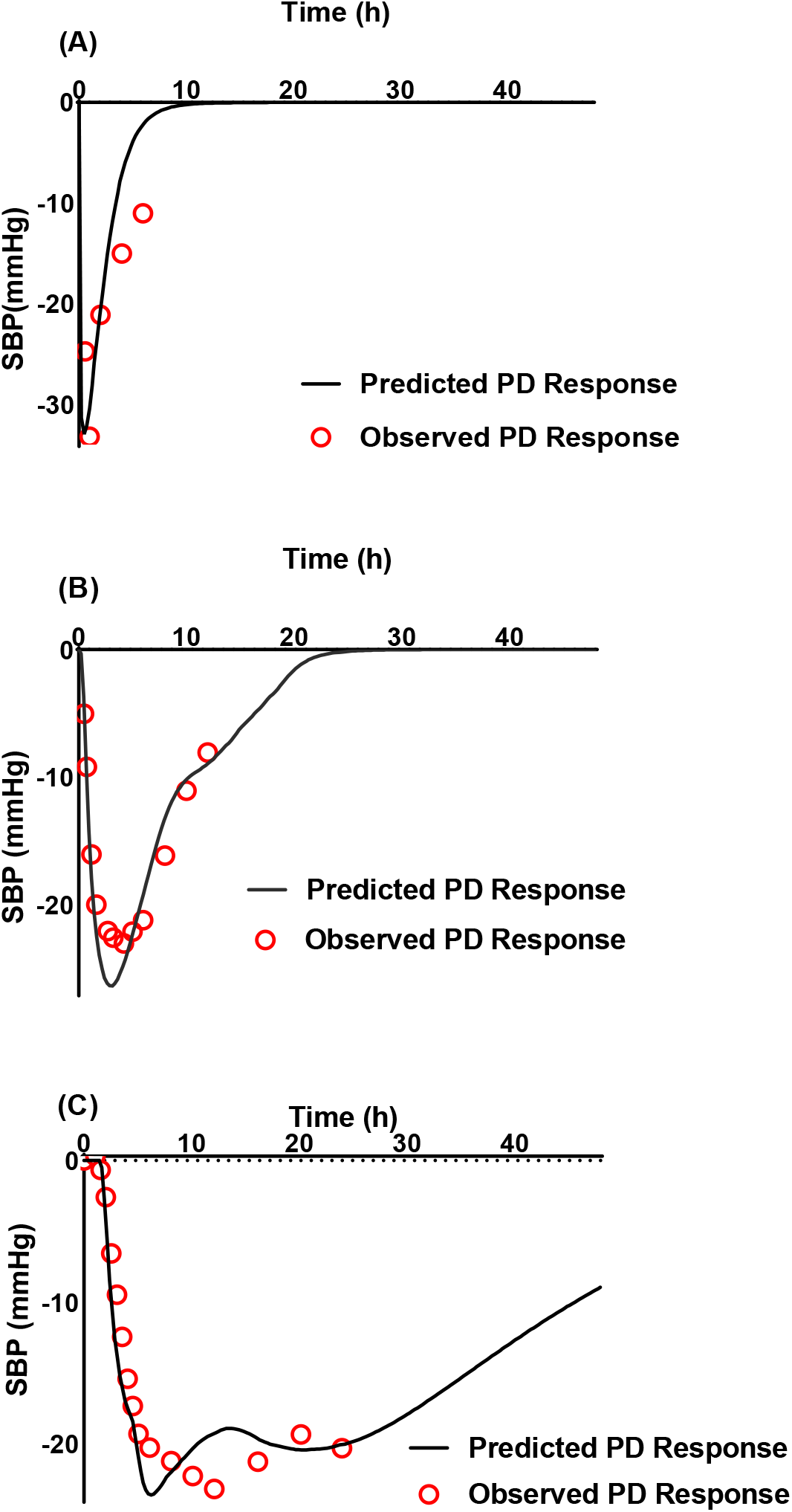
Comparison of model predicted systolic blood pressure (SBP) compared with clinical observations in patients after a single dose of (A) 10 mg IR nifedipine; (B) 20mg SR nifedipine; (C) 60 mg CR nifedipine. Red circles represent observed values and black line represent predicted values.

**Table 7.** Comparison of the predicted and observed maximum reduction in systolic blood pressure (R_max_) and the area under the effect-time curve (AUE) for the different nifedipine formulations.

|  | Immediate-Release (IR) nifedipine |  |  | Sustained-Release (SR) nifedipine |  |  | Controlled-Release (CR) nifedipine |  |  |
| --- | --- | --- | --- | --- | --- | --- | --- | --- | --- |
|  | Pred | Obs | Fold error | Pred | Obs | Fold error | Pred | Obs | Fold error |
| $R_{\max}$ (mmHg) | -32.93 | -33.2 | 0.99 | -26.35 | -23.04 | 1.14 | -23.61 | -23.18 | 1.02 |
| AUE (mmHg·h) | 88.33 | 102 | 0.87 | 196.6 | 195 | 1.01 | 428.4 | 435.4 | 0.98 |

*Obs*, observation; *Pred*, prediction, *Fold-error*, ratio of predicted: observed values.

### Nifedipine-RTV PBPK/PD model application

For the three formulations of nifedipine, dose adjustment of nifedipine during RTV co-administration was unable to maintain nifedipine plasma concentrations and SBP at the same level as without RTV (**Figure 8**). Moreover, the established PBPK model showed that reducing the daily dose of IR nifedipine from 20 to 1.25 mg did not significantly alter the nifedipine–RTV DDI potential, suggesting that reduced-dose (1/16) nifedipine (**Figure S1**) might not fully mitigate the risk of severe hypotension when combined with standard-dose (100 mg) RTV. The influence of dose reduction on the SR and CR formulations are the same as IR.

**Figure 8.**
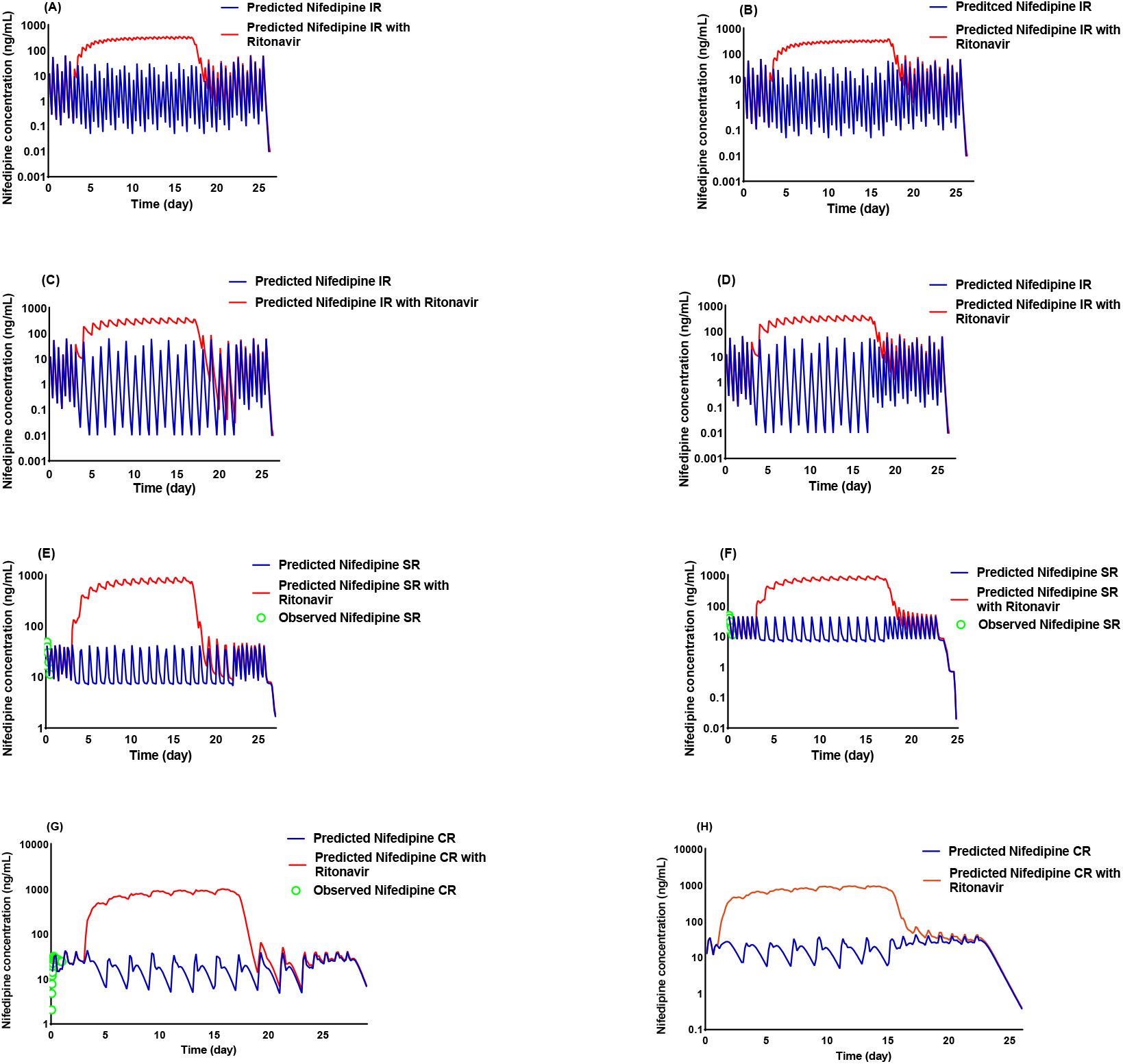
PK simulation results of nifedipine dose adjustment in different scenarios after RTV discontinuation. (A) IR nifedipine 10 mg every 12 hours (Q12H) × 3 days + (5 mg Q12H + RTV 100 mg Q12H) × 14 days+ nifedipine 5 mg Q12H x5 days + nifedipine 10 mg Q12H × 1 day. (B) IR nifedipine 10 mg Q12H × 3 days + (5 mg Q12H + RTV 100 mg Q12H) x14 days+ nifedipine 10 mg Q12H × 6 days. (C) IR nifedipine 10 mg Q12H × 3 days + (10 mg Q24H + RTV 100 mg Q12H) x14 days + nifedipine 10 mg Q24H × 5 days + nifedipine 10 mg Q12H × 1 day. (D) IR nifedipine 10 mg Q12H × 3 days + (10 mg Q24H + RTV 100 mg Q12H) x14 days+ nifedipine 10 mg Q12H × 6 days. (E) SR nifedipine 30 mg Q12H × 3 days + (30 mg Q24H + RTV 100 mg Q12H) × 14 days + nifedipine 30 mg Q24H × 5 days + nifedipine 30 mg Q12H × 1 day. (F) SR nifedipine 30mg Q12H × 3 days + (30 mg Q24H + RTV 100 mg Q12H) × 14 days + nifedipine 30 mg Q12H × 6 day. (G) CR nifedipine 60 mg Q24H × 3 days + (60 mg Q48H + RTV 100 mg Q12H) × 14 days + nifedipine 60 mg Q24H × 5 days + nifedipine 60 mg Q24H × 1 day. (H) CR nifedipine 60 mg Q24H × 3 days + (60 mg Q48H + RTV 100 mg Q12H) x14 days + nifedipine 60 mg Q24H × 6 days. Red and blue lines represent predicted values for nifedipine with and without co-administration of RTV, respectively.

For scenario 1, **Figure 9** shows the PD profiles of regular dose of nifedipine combined with RTV. For an individual taking a regular nifedipine dose combined with 100 mg Q12H RTV, the predicted dynamic SBP decrease was up to 47 mmHg, which might be a critically low blood pressure. Moreover, nifedipine at a reduced dose during RTV co-administration was unable to maintain SBP in normal range (**Figure 10**). Therefore, the combined use of nifedipine and RTV-containing regimens is not recommended.

**Figure 9.**
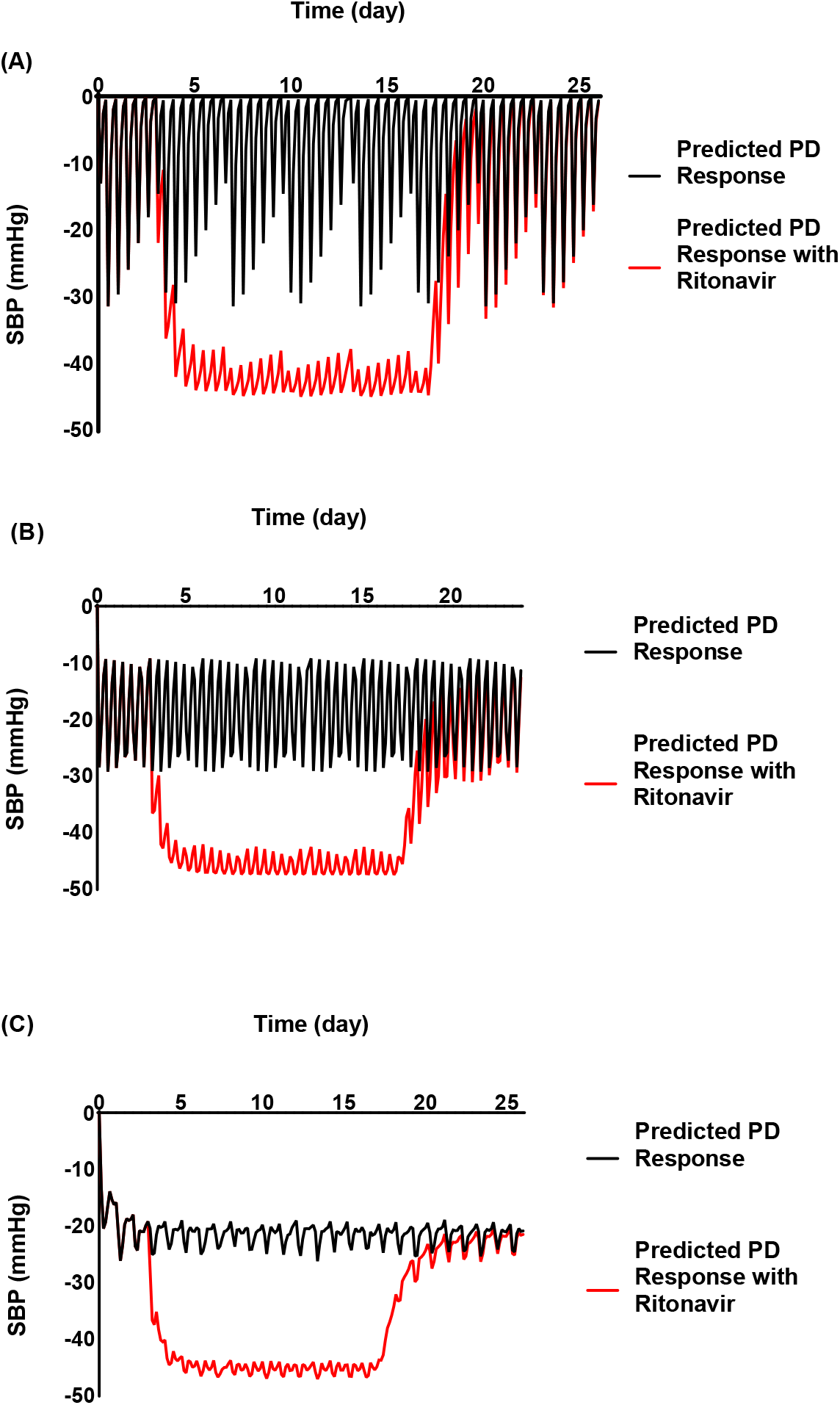
Model prediction of SBP changes in the drug–drug interaction (DDI) magnitude of nifedipine and ritonavir (RTV) over multiple days. (A) IR nifedipine 10 mg every 12 hours (Q12H) × 3 days + (nifedipine 10 mg Q12H + RTV 100 mg Q12H) × 14days+ nifedipine 10 mg Q12H × 10 days. (B) SR nifedipine 30 mg Q12H × 3 days + (30 mg Q12H +RTV 100 mg Q12H) x14 days+ nifedipine 30 mg Q12H × 10 days. (C) CR nifedipine 60 mg Q24H × 3 days+ (60 mg Q24H + RTV 100 mg Q12H) x14 days+ nifedipine 60 mg Q24H × 10 days. The red and black solid lines represent the PD curves of combined nifedipine with RTV and nifedipine alone, respectively.

**Figure 10.**
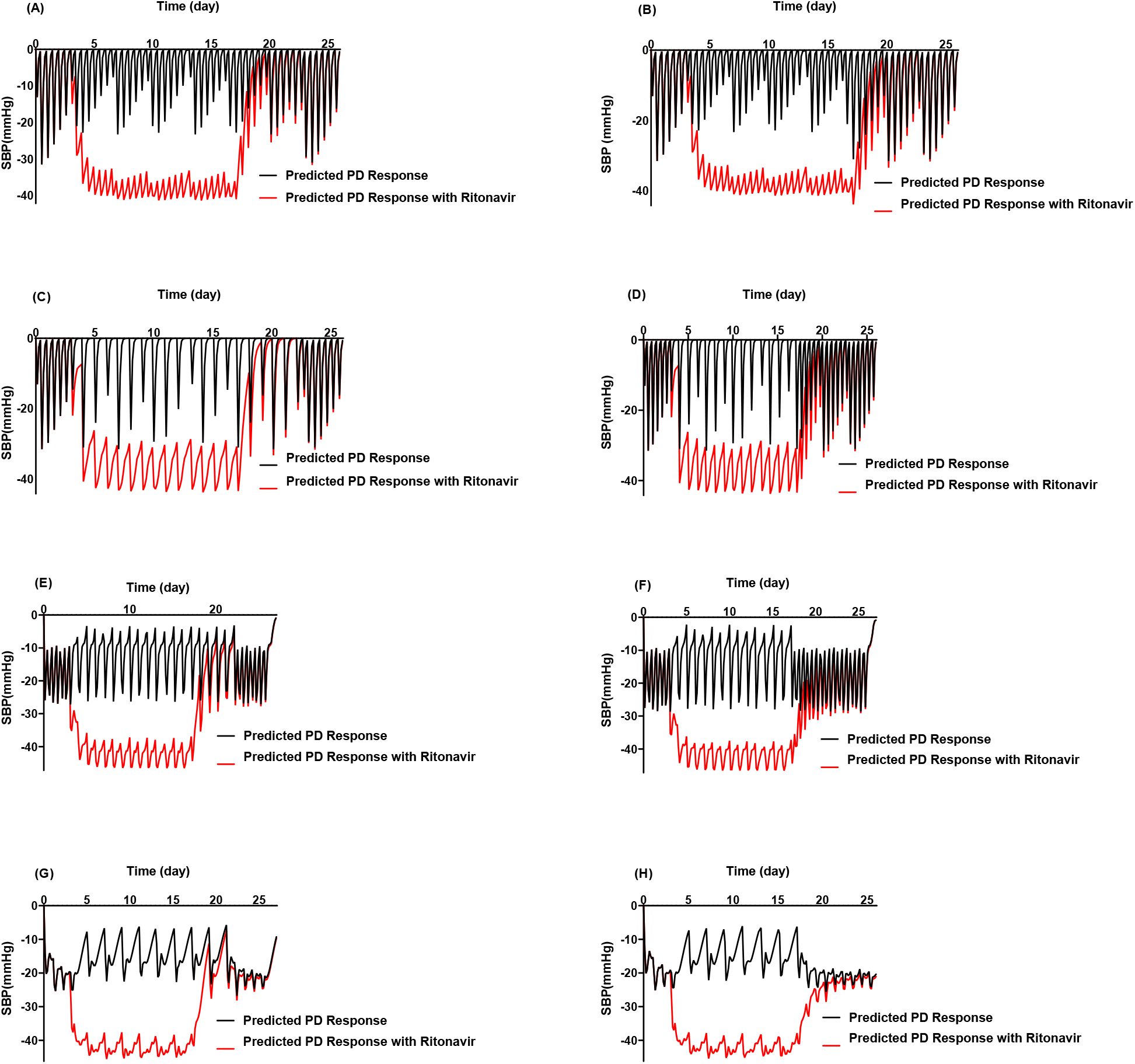
PD simulation results of nifedipine dose adjustment in different scenarios after RTV discontinuation. (A) IR nifedipine 10 mg every 12 hours (Q12H) × 3 days + (5 mg Q12H + RTV 100 mg Q12H) × 14 days+ nifedipine 5 mg Q12H × 5 days + nifedipine 10 mg Q12H × 1 day. (B) IR nifedipine 10 mg Q12H × 3 days + (5 mg Q12H + RTV 100 mg Q12H) × 14 days+ nifedipine 10 mg Q12H × 6 days. (C) IR nifedipine 10 mg Q12H × 3 days + (10 mg Q24H + RTV 100 mg Q12H) x14 days + nifedipine 10 mg Q24H × 5 days + nifedipine 10 mg Q12H × 1 day. (D) IR nifedipine 10 mg Q12H × 3 days + (10 mg Q24H + RTV 100 mg Q12H) x14 days+ nifedipine 10 mg Q12H × 6 days. (E) SR nifedipine 30 mg Q12H × 3 days + (30 mg Q24H + RTV 100 mg Q12H) × 14 days + nifedipine 30 mg Q24H × 5 days + nifedipine 30 mg Q12H × 1 day. (F) SR nifedipine 30mg Q12H × 3 days + (30 mg Q24H + RTV 100 mg Q12H) × 14 days + nifedipine 30 mg Q12H × 6 day. (G) CR nifedipine 60 mg Q24H × 3 days + (60 mg Q48H + RTV 100 mg Q12H) × 14 days + nifedipine 60 mg Q24H × 5 days + nifedipine 60 mg Q24H × 1 day. (H) CR nifedipine 60 mg Q24H × 3 days + (60 mg Q48H + RTV 100 mg Q12H) x14 days + nifedipine 60 mg Q24H × 6 days. The open square symbols correspond to the mean observed values. Red and black lines represent predicted values for nifedipine with and without co-administration of RTV, respectively.

The simulations from scenarios 2 (**Figure 10**) showed continuing the reduced nifedipine dose for an additional 5 days results in a lower nifedipine plasma concentrations and a corresponding increase in SBP over the 5 days. This suggested that the dose of nifedipine cannot be immediately restored to the regular dose after the withdrawal of RTV in case of RTV co-administration.

## Discussion

With the outbreak of viral infections such as COVD-19, MERS and SARS, the antiviral effect of RTV-containing regimens has received increasing attention [16, 45]. Hypertensive patients are often potential susceptible population [46, 47] and require antiviral treatment after viral infection. Thus, the combination of CCB and RTV is not unavoidable. There have been previous studies [35, 48] on the DDI of RTV and amlodipine, but there is a lack of systematic research on the commonly used CCB, nifedipine. To the best of our knowledge, this is the first systematic study to investigate the DDI between nifedipine and RTV-containing regimens by using PBPK/PD analysis.

Previously published PBPK modeling of nifedipine mainly focused on the drug formulations [23, 49] or special populations [50-52]. The nifedipine model developed in this study is more comprehensive considering commonly used three formulations, relationship of PK and PD and DDI. Moreover, the nifedipine PBPK model built in the software of Simcyp® hasn’t been well verified for the purpose of DDI prediction. The developed nifedipine model was herein verified with clinical DDI studies involving CYP3A inhibitors/inducers or with PK profiles from the healthy volunteers. Although the absolute values of the model-predicted C_max_ and AUC did not match the observed values perfectly, the exposure change caused by DDI were in good agreement with the observed data (all the fold errors <1.83) (**Table 4**). This indicates the good performance of nifedipine PBPK model as a victim drug in the DDI prediction.

Nifedipine undergoes significant first-pass metabolism by CYP3A in the both intestine and liver[53], thus resulting in significantly enhanced *in vivo* exposure of the drug when administered together with strong irreversible CYP3A4 inhibitor like RTV. The inhibitory potency of RTV *in vivo* has been shown to be equivalent to or greater than ketoconazole which is a strong index CYP3A inhibitor for DDI studies [54]. The PBPK model described the interaction over time between nifedipine and RTV, and showed that the combined use of RTV significantly reduced the CYP3A4 enzyme content in the liver and intestine and the C_max_ and AUC_24h_ of nifedipine increased by 9.82-34.35 and 44.94-50.77 times, respectively. The C_max_ of all three formulations nifedipine exceeded 700 ng/ml, which is far beyond the therapeutic concentration range of nifedipine (25-100 ng/mL)[6]. Therefore, the potential risk of severe hypotension becomes very high after its combined use with RTV.

Moreover, the established PBPK/PD model showed that nifedipine dose reduced to 1/16 of the regular dose during RTV co-administration couldn’t prevent the risk of hypotension. Therefore, patients are not recommended to take any formulations of nifedipine and RTV-containing regimens simultaneously. In addition, it takes 5 days of wash-out after RTV withdrawal to allow the nifedipine concentration to drop down to a safe level when patient taking nifedipine. These results are the important hints for patients taking the nifedipine treatment.

The PBPK/PD analysis was once used to investigate dose adjustment recommendations for amlodipine during and after co-administration of RTV by Mukherjee, *et al* [35]. The analysis suggested that resuming a full dose of amlodipine (5 mg QD) immediately or continuing with the reduced dose (2.5 mg QD) for 5 days after the last dose of RTV could be appropriate. Based on the simulation of this study, the effect of RTV on nifedipine PK is significantly stronger than that on amlodipine, although nifedipine and amlodipine are both dihydropyridine CCBs. Compared with nifedipine, amlodipine has a lower incidence of interactions due to less first-pass metabolism[55]by CYP3A isoform in the intestine and liver.

This study systematically investigated the DDI between nifedipine and RTV-containing regimens, and provided meaningful guidance for clinical use, especially during the COVID-19 pandemic. The recent open-label, randomized, phase 2 trial in patients with COVID-19 showed that triple combinations of interferon beta-1b, LPV/r and ribavirin was safe and superior to LPV/r alone in alleviating symptoms and shortening the duration of viral shedding and hospital stay in patients with mild to moderate COVID-19[17]. And it’s reported that among 5700 patients hospitalized with COVID-19 in the New York City area, even up to 56.6% have comorbid hypertension [56]. Thus, the scenario of combining RTV during nifedipine drug treatment would probably be of great clinical relevance. Our study showed that it could lead to severer hypotension for patients with COVID-19 to take nifedipine and RTV-containing regimens simultaneously.

Due to the lack of pharmacodynamic studies of nifedipine overdose, PD model in this study referred to the previously reported model and verified with clinical data at therapeutic doses, which may not accurately predict blood pressure changes at excessive dose. However, the exposure of nifedipine regardless of the formulation has increased many times after the combined use of RTV, far beyond the normal range. Therefore, the combined use of nifedipine and RTV-containing regimens is still not recommended according to PK prediction results. If an antiviral regimen containing RTV is required, other antihypertensive agents should be replaced.

## Conclusions

RTV had a pronounced effect on the pharmacokinetics of nifedipine. Combinations of nifedipine and RTV is not recommended according to the PBPK/PD analysis. Restart of nifedipine 5 days after discontinuation of RTV can maintain plasma levels and blood pressure at a relatively safe level.

## Data Availability

All data generated or analyzed during this study are included in this article.

---

**Supplementary Figure 1.**
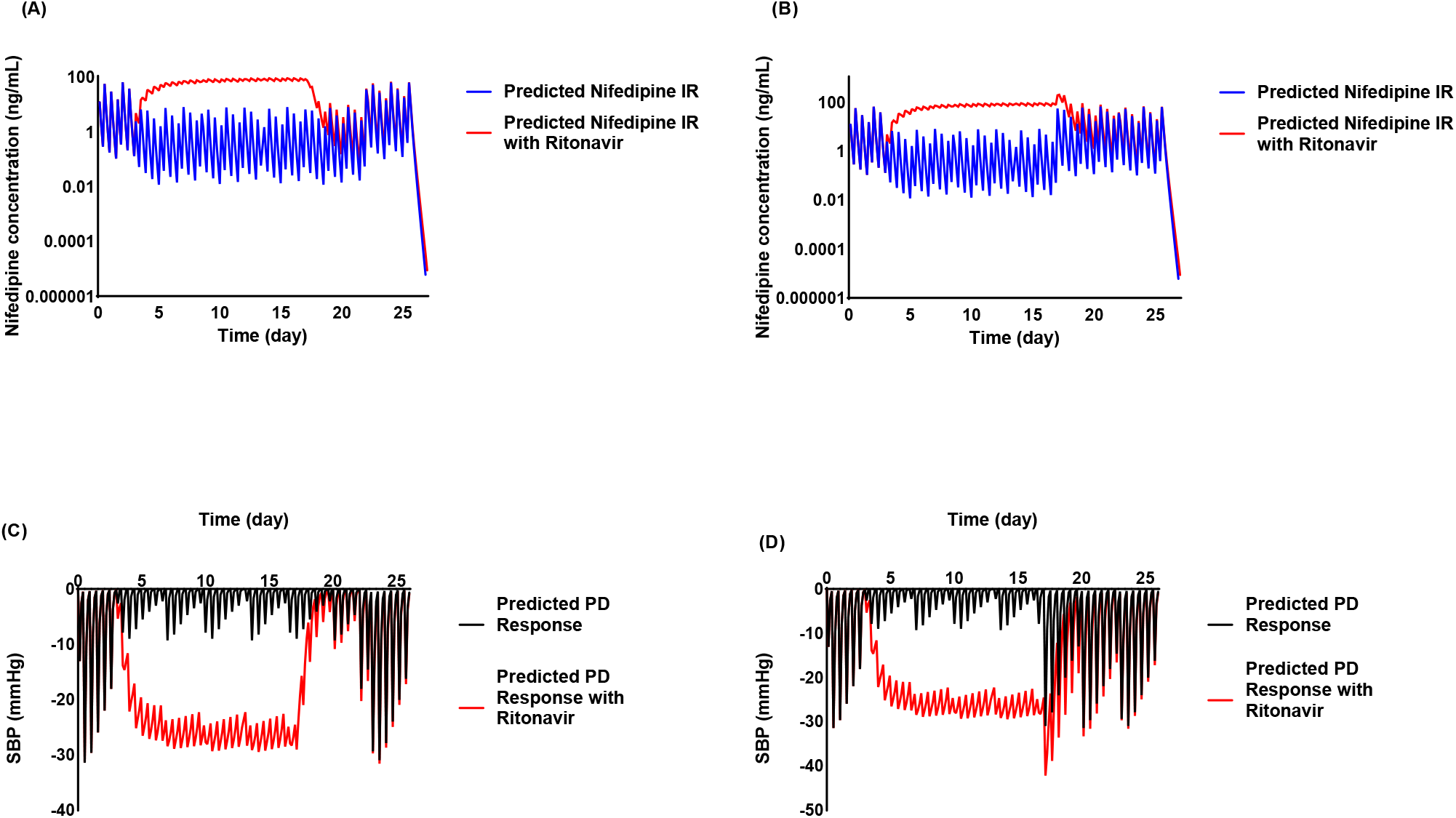
PK (A, B) and PD (C, D) simulation results of IR nifedipine dose adjustment in following scenarios after RTV discontinuation. (A)(C)10 mg Q12H × 3 days + (1.25 mg Q12H + RTV 100 mg Q12H) × 14 days + 1.25 mg Q12H × 5 days + 10 mg Q12H; (B)(D)10 mg Q12H × 3 days + (1.25 mg Q24H + RTV 100 mg Q12H) × 14 days + 1.25 mg Q24H × 5 days + 10 mg Q12H.

## Author contribution

**Zheng Jiao:** Conceptualization, Writing-Original draft; Writing-Review & Editing; Supervision; Project administration; Funding acquisition. **Xiao-qiang Xiang:** Methodology; Validation; Resources; Writing-Original draft; Writing-Review & Editing; Supervision; Project administration; Funding acquisition. **Si-ze Li**: Formal analysis; Software; Investigation; Data Curation. **Xi-ying Lin**: Investigation, Formal analysis. **Sha-sha Jin**: Investigation; Visualization. **Meng-wan Zhang**: Data Curation, Funding acquisition. **Ming-kang Zhong** and **Wei-min Cai**: Supervision. **Wan-jie Niu**: Investigation; Writing-Original draft; Writing-Review & Editing; Visualization.

